# Insecticide resistance outpaces behavioural adaptation, as a response to Long-Lasting Insecticidal Net distribution, in malaria vectors in Burkina Faso

**DOI:** 10.1101/2021.06.01.21258151

**Authors:** Antoine Sanou, Luca Nelli, W. Moussa Guelbéogo, Fatoumata Cissé, Michel M. Tapsoba, Pierre Ouédraogo, N’falé Sagnon, Hilary Ranson, Jason Matthiopoulos, Heather M. Ferguson

## Abstract

The decline in malaria across Africa has been largely attributed to vector control using Long-Lasting Insecticidal Nets (LLINs). However, this intervention has prompted widespread insecticide resistance (IR) and been associated with changes in mosquito behaviour that reduce their contact with LLINs. The relative importance and rate at which IR and behavioural adaptations emerge are poorly understood. We conducted surveillance of mosquito behaviour and IR at 12 sites in Burkina Faso to assess the magnitude and temporal dynamics of insecticide and behavioural resistance in vectors in the 2-year following mass LLIN distribution. Insecticide resistance was present in all vector populations and increased rapidly over the study. In contrast, no longitudinal shifts in LLIN-avoidance behaviours (earlier or outdoor biting and resting) were detected. There was a moderate shift in vector species composition from *Anopheles coluzzii* to *Anopheles gambiae* which coincided with a reduction in the proportion of bites preventable by LLINs; possibly driven by between-species variation in behaviour. These findings indicate that adaptations based on insecticide resistance arise and intensify more rapidly than behavioural shifts within mosquito vectors. However, longitudinal shifts in mosquito vector species composition were evident within 2 years following a mass LLIN distribution. This ecological shift was characterized by a relative increase in the moderately more exophagic species (*An. gambiae*) and coincided with a predicted decline in the degree of protection expected from LLINs. Although human exposure fell through the study period due to reducing vector densities and infection rates, such ecological shifts in vector species along with insecticide resistance were likely to have eroded the efficacy of LLINs. While both adaptations impact malaria control, the rapid increase of the former indicates it is the most rapid strategy but interventions targeting both will be needed.

## Introduction

Long-lasting Insecticidal nets (LLINs) and Indoor Residual Spraying (IRS) are the main malaria vector controls tools [1, 2] recommended by the World Health Organisation (WHO) [3]. Both interventions have had a significant impact on malaria control in Africa; with LLINs being primarily responsible for the >50% reduction in prevalence since 2000 [4]. The success of these interventions rely on their ability to exploit innate aspects of malaria vector behaviour including a preference for feeding on humans (anthropophagy), inside houses (endophagy), during sleeping hours, and to rest inside houses (endophily) after feeding [5, 6]. Consequently, LLINs and IRS are expected to work best against anthropophilic, endophilic and endophagic vectors that have high susceptibility to insecticides [7, 8]. Historically, most of the major malaria vectors in Africa (members of the *An. gambiae* s.l. complex) were described as having highly anthropophilic [9, 10] and endophilic resting behaviour [11], characterized by late night-biting [11, 12]. This combination of traits is thought to account for the early success of current vector control approaches in Africa [13].

However global progress on malaria control has stalled since 2015 [14], with a slowing of decline in cases and deaths, and even an increase in some settings. This stagnation has been most pronounced within a small group of high-burden countries including ten in Africa plus India. This slowdown is hypothesized to be driven by the widespread emergence of insecticide resistance (IR) in malaria vectors that has now been documented in almost all African countries [15]. Resistance has been facilitated by the reliance of almost all LLIN products on a single class of insecticides (pyrethroids). Vectors have developed a range of resistance mechanisms against pyrethroids including. target site mutations [16], cuticular resistance [17] and metabolic resistance [18]. Additionally, there are reports of vectors shifting key behaviours that predispose them to LLINs including their host choice [19], biting time [7, 20] and location (indoor versus outdoors, [21]) and resting [22]. Such shifts can arise either through ecological changes in vector species composition (with more zoophilic, exophagic and exophilic species being favoured; [23, 24]) or evolutionary adaptations reflected by behavioural changes within species. Examples of within species behavioural changes include the apparent switch in biting time from late night to morning in *An funestus* in Benin [25] and from indoor to outdoor resting in *An. arabiensis* in Tanzania [19]. Such ecological and adaptive changes have been documented in several settings [26, 27]. Regardless of how changes in vector behaviour arise, they are expected to erode LLIN effectiveness, with outdoor biting alone estimated as being responsible for over 10 million malaria cases in Africa [28].

Ecological shifts and evolutionary adaptations in mosquito behaviour could both reduce malaria control by enabling mosquitoes to avoid contact with LLINs. In contrast, IR allows mosquitoes to tolerate contact with LLINs by avoiding immediate mortality. Understanding the relative capacity of mosquito vectors to mount IR and behavioural avoidance in response to LLINs is necessary to estimate the risk posed by each process to malaria control and highlight where mitigation efforts should be prioritized. For instance, rapid emergence of behavioural avoidance mandates incorporation of interventions that target mosquitoes outside homes (e.g. spatial repellents, endectocides, larvicides etc [29]). In contrast, the rapid emergence of insecticide resistance in vectors who continue to feed and rest indoors indicates that switching to alternative insecticide classes in indoor-based interventions (e.g. next generation nets or using non-insecticidal methods inside the home (house screening [30, 31]) may have greater impact.

Burkina Faso is amongst the 10 high burden malaria countries in Africa where prevalence appears to be rising despite several rounds of mass LLIN distribution [32]. The *Anopheles gambiae* (*An. arabiensis, An. coluzzii* and *An. gambiae)* and *An. funestus* species groups are the primary vectors in this country [33, 34]. Resistance to insecticides (DDT and Dieldrin) was first detected in Burkina Faso in the late 1960s [35, 36]. Over the past two decades IR has intensified in vector populations in Burkina Faso [15, 37] with levels being amongst the highest in Africa [38]. This high rate of IR is hypothesized to be responsible for the limited impact of LLINs in Burkina Faso [39], but less is known about the potential of additional impacts of mosquito behavioural adaptations and changes in vector species composition. In the current study we estimated the potential epidemiological impact of temporal shifts in vector abundance, insecticide resistance and behaviour in two ways. First, we estimated expected human exposure to infected mosquitoes in term of the “Entomological Inoculation Rate “(“EIR”) between study years. The EIR is considered to be one of the most direct estimates of human exposure to malaria and has a relatively good correlation with human epidemiological outcomes such as malaria incidence [40, 41]. Additionally, we estimated the degree of protection expected to be obtained from use of an effective LLIN during typical sleeping hours, as a function of the time and location of mosquito biting [42]. In combination this information will enable assessment of malaria transmission and the expected impact of current control tools as vector behavioural and physiological resistance rises.

Much of the knowledge on malaria vector behaviours in Burkina Faso comes from before the period of mass LLIN distribution [43-45] and showed that that > 50% of *An. gambiae* s.l. bite indoors and biting activity was concentrated late at night (01 h – 06 h), with only few studies thereafter [46, 47]. We conducted an observational study to measure the intensity and rate of change in insecticide and behavioural resistance traits within malaria vector populations over 2 years following a mass LLIN distribution in Burkina Faso. Aims were to (1) update information on vector ecology and behaviour in a high burden African country, (2) quantify and compare the magnitude and rate of change in insecticide and behavioural resistance traits, and malaria vector species composition and (3) estimate the potential impact of changes in these ecological and physiological traits on malaria transmission and LLIN effectiveness

## Results

A total of 49,482 mosquitoes (both female and male) were caught in HLC, of which most were female *An. gambiae* s.l. (N = 40,220; Table 1). A total of 927 mosquitoes were collected from the RBTs of which 584 were female *An. gambiae* s.l. (Table 2). Within the subset (∼20 % of total) of *An. gambiae* s.l. collected in HLC that were identified to species level, 53.2% were *An. coluzzii*, 45.9% were *An. gambiae* and 0.28% were *An. arabiensis*. Within the 449 resting female *An. gambiae* s.l. identified by PCR, *An. coluzzii* was dominant (61.25%) followed by *An. gambiae* (38.08%), *An. arabiensis* (n=2, 0.45%) and one hybrid between *An. coluzzii* and *An. gambiae* (0.22). Due to the low number of *An. arabiensis*, this species was not included in statistical analysis of species-specific differences.

**Table 1:** Total number of female Anophelines caught using Human Landing Catches in 12 villages of southwestern Burkina Faso, from October 2016 to December 2018. Results are displayed by trapping location (IN = indoor and OUT = outdoor) and village.

|  | *An. coustani* |  | *An. funestus* |  | *An. gambiae s.l.* |  | *An. nili* |  | *An. obscurus* |  | *An. pharoensis* |  | *An. rufipes* |  | *An. zeimani* |  | Total |
| --- | --- | --- | --- | --- | --- | --- | --- | --- | --- | --- | --- | --- | --- | --- | --- | --- | --- |
| Village | IN | OUT | IN | OUT | IN | OUT | IN | OUT | IN | OUT | IN | OUT | IN | OUT | IN | OUT |  |
| Dangouindougou | 0 | 4 | 3 | 3 | 788 | 786 | 0 | 1 | 0 | 0 | 11 | 14 | 1 | 0 | 0 | 0 | 1,611 |
| Gouera | 0 | 0 | 0 | 5 | 762 | 867 | 0 | 3 | 0 | 0 | 1 | 4 | 0 | 0 | 0 | 0 | 1,642 |
| Nianiagara | 0 | 0 | 1 | 0 | 451 | 522 | 4 | 0 | 0 | 0 | 2 | 4 | 0 | 0 | 0 | 0 | 984 |
| Nofesso | 0 | 0 | 1 | 0 | 340 | 540 | 0 | 0 | 0 | 0 | 0 | 0 | 0 | 0 | 0 | 0 | 881 |
| Ouangolodougou | 0 | 0 | 0 | 0 | 265 | 409 | 0 | 0 | 0 | 0 | 1 | 0 | 0 | 0 | 0 | 0 | 675 |
| Sitiena | 31 | 12 | 1 | 1 | 2,549 | 2,589 | 160 | 306 | 5 | 8 | 25 | 34 | 0 | 0 | 1 | 0 | 5,722 |
| Tengrela | 49 | 88 | 4 | 3 | 4,579 | 4,598 | 25 | 47 | 1 | 0 | 63 | 152 | 2 | 5 | 2 | 3 | 9,621 |
| Tiefora | 1 | 2 | 4 | 2 | 3,917 | 4,754 | 14 | 24 | 0 | 0 | 52 | 119 | 0 | 0 | 0 | 0 | 8,889 |
| Timperba | 0 | 1 | 0 | 1 | 445 | 413 | 0 | 1 | 0 | 0 | 0 | 0 | 0 | 0 | 0 | 0 | 861 |
| Tondoura | 0 | 0 | 1 | 0 | 1,121 | 1,096 | 2 | 5 | 0 | 0 | 1 | 1 | 0 | 0 | 0 | 0 | 2,227 |
| Toumousseni | 0 | 2 | 2 | 2 | 2,670 | 3,056 | 8 | 17 | 0 | 0 | 14 | 29 | 0 | 0 | 0 | 0 | 5,800 |
| Yendere | 2 | 1 | 0 | 4 | 1,272 | 1,431 | 5 | 9 | 0 | 0 | 9 | 9 | 0 | 0 | 0 | 0 | 2,742 |
| Total | 83 | 110 | 17 | 21 | 19,159 | 21 061 | 218 | 413 | 6 | 8 | 179 | 366 | 3 | 5 | 3 | 3 | 41,655 |

**Table 2:**
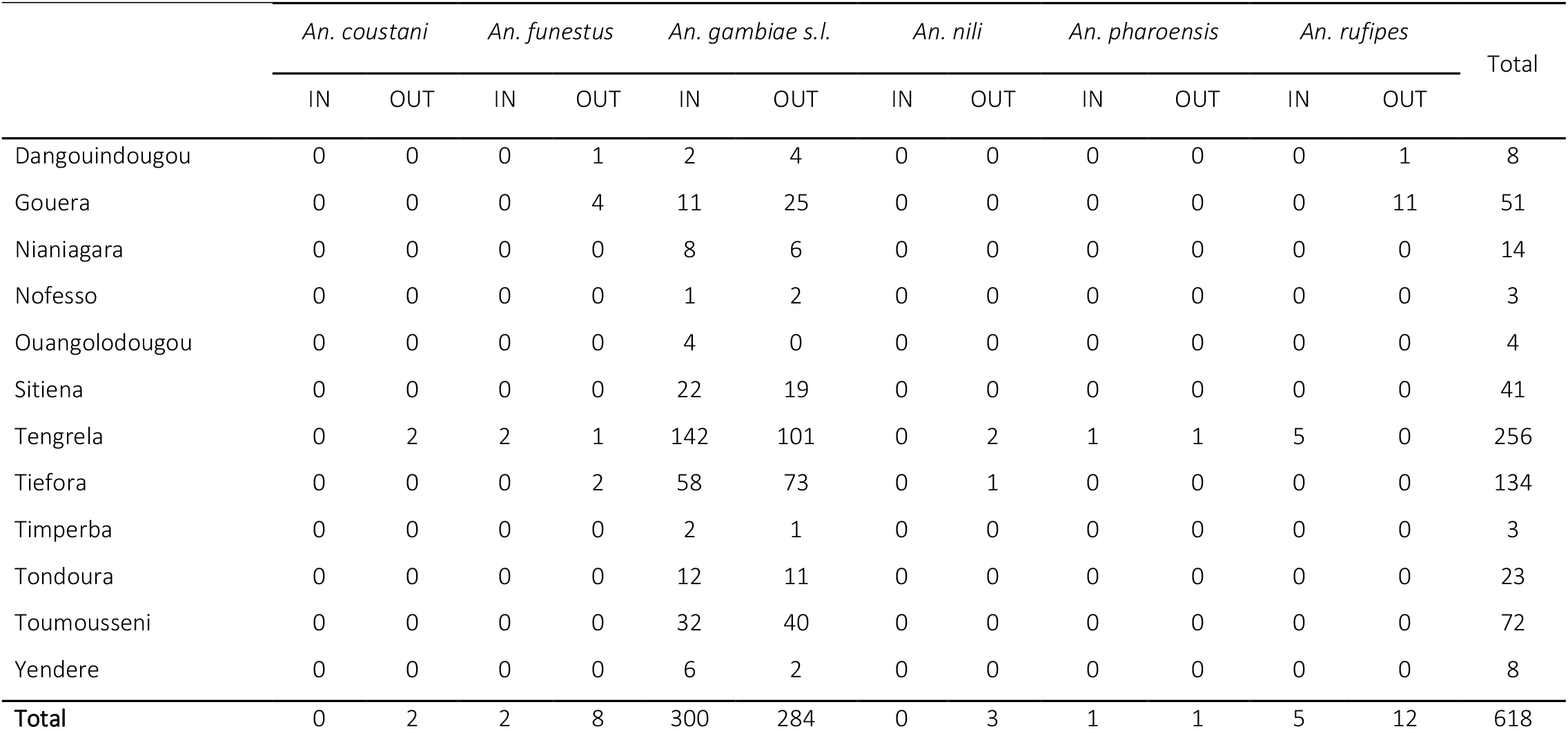
Total number of female Anophelines caught using Resting Bucket Traps in 12 villages of southwestern Burkina Faso, from October 2016 to December 2018. Results are displayed by trapping location (IN = indoor and OUT = outdoor) and village.

|  | *An. coustani* |  | *An. funestus* |  | *An. gambiae s.l.* |  | *An. nili* |  | *An. pharoensis* |  | *An. rufipes* |  | Total |
| --- | --- | --- | --- | --- | --- | --- | --- | --- | --- | --- | --- | --- | --- |
|  | IN | OUT | IN | OUT | IN | OUT | IN | OUT | IN | OUT | IN | OUT |  |
| Dangouindougou | 0 | 0 | 0 | 1 | 2 | 4 | 0 | 0 | 0 | 0 | 0 | 1 | 8 |
| Gouera | 0 | 0 | 0 | 4 | 11 | 25 | 0 | 0 | 0 | 0 | 0 | 11 | 51 |
| Nianiagara | 0 | 0 | 0 | 0 | 8 | 6 | 0 | 0 | 0 | 0 | 0 | 0 | 14 |
| Nofesso | 0 | 0 | 0 | 0 | 1 | 2 | 0 | 0 | 0 | 0 | 0 | 0 | 3 |
| Ouangolodougou | 0 | 0 | 0 | 0 | 4 | 0 | 0 | 0 | 0 | 0 | 0 | 0 | 4 |
| Sitiena | 0 | 0 | 0 | 0 | 22 | 19 | 0 | 0 | 0 | 0 | 0 | 0 | 41 |
| Tengrela | 0 | 2 | 2 | 1 | 142 | 101 | 0 | 2 | 1 | 1 | 5 | 0 | 256 |
| Tiefora | 0 | 0 | 0 | 2 | 58 | 73 | 0 | 1 | 0 | 0 | 0 | 0 | 134 |
| Timperba | 0 | 0 | 0 | 0 | 2 | 1 | 0 | 0 | 0 | 0 | 0 | 0 | 3 |
| Tondoura | 0 | 0 | 0 | 0 | 12 | 11 | 0 | 0 | 0 | 0 | 0 | 0 | 23 |
| Toumousseni | 0 | 0 | 0 | 0 | 32 | 40 | 0 | 0 | 0 | 0 | 0 | 0 | 72 |
| Yendere | 0 | 0 | 0 | 0 | 6 | 2 | 0 | 0 | 0 | 0 | 0 | 0 | 8 |
| **Total** | 0 | 2 | 2 | 8 | 300 | 284 | 0 | 3 | 1 | 1 | 5 | 12 | 618 |

### Insecticide Resistance

The overall mortality of *An. gambiae* s.l. in the 24 hours following exposure to the DD was 23.33% [95%CI: 14.63 – 32.05%], indicating a resistant population according to WHO criteria. The mortality of *An. gambiae* s.l. following exposure to the DD of deltamethrin declined significantly over the study period, falling from ∼38% at the beginning (October 2016) to ∼17% toward the end (September 2018, Figure 1). As expected, mortality 24 hour post-exposure generally rose with increased concentration of deltamethrin ((0.25%: 64.14% [95% CI: 55.27 – 73.0]; 0.5%: 86.03 [95% CI: 81.87 – 90.2%]; 0.75%: 87.48% [95% CI: 81.52 – 92.43%]). The predicted reduction in post-exposure mortality over time as observed with the DD was also detected at 15X the DD (Supplementary Table 1). Long-term increases in IR were evident after controlling for statistically significant variation between villages (all concentrations) and season (only detected in bioassays using the DD; Supplementary Table 1).

**Figure 1:**
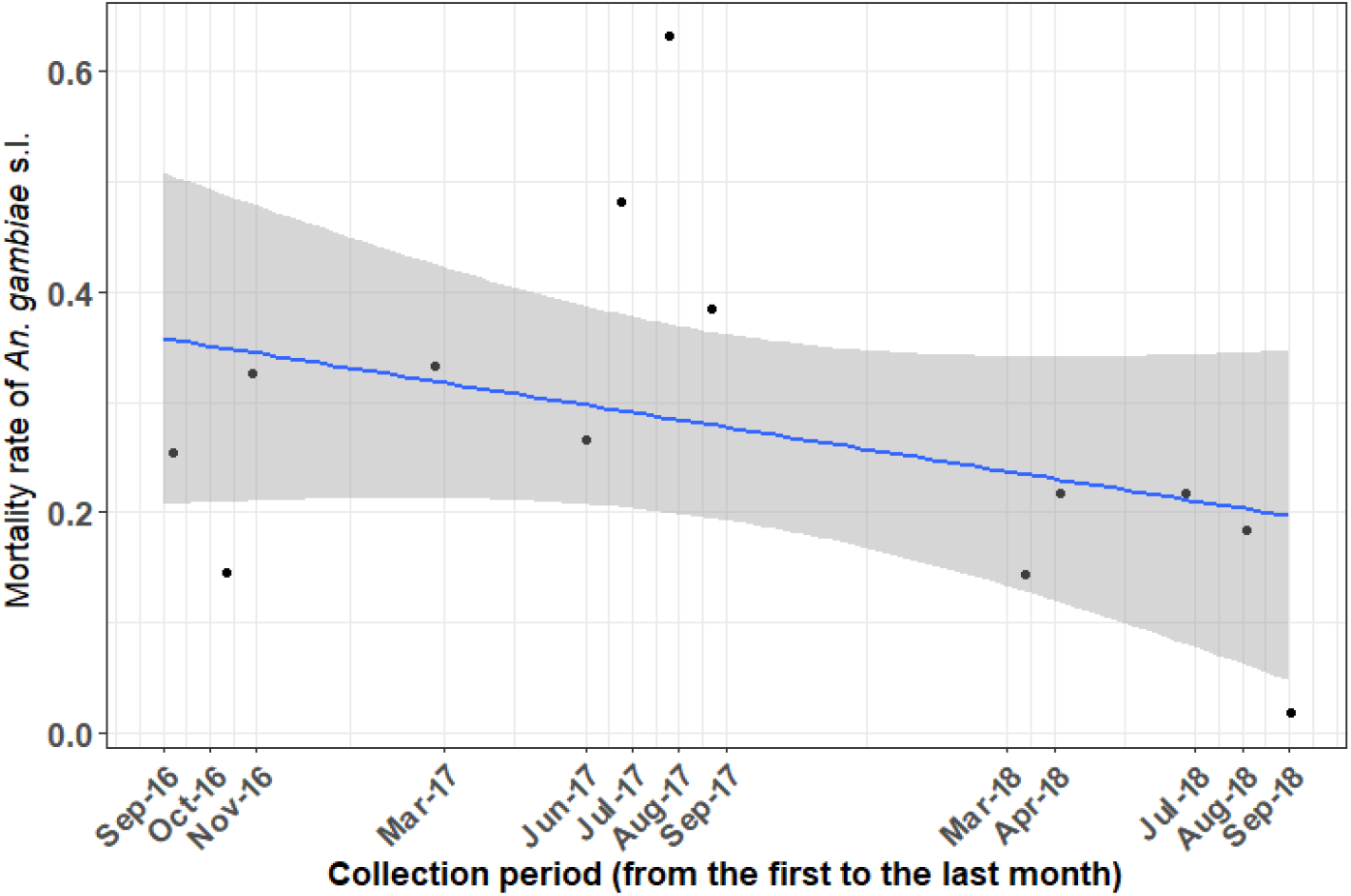
Mean predicted longer-term trend in the mortality of *Anopheles gambia*e s.l. 24 hours following exposure to a diagnostic dose of deltamethrin across the study period. Black dots indicate mean predicted mortality at each study month between September 2016 to September 2018 across 9 villages in southwestern Burkina Faso. The blue line indicates the predicted linear change in *An. gambiae* s.l. across the study period based on the final model, with the grey-shaded areas around them indicate 95% confidence intervals.

### The human biting rate and *Anopheles gambiae* s.l. species composition

Across the whole study period and pooling across sites, the mean HBR of *An. gambiae* s.l. was ∼17 bites per person, per night (bppn). After controlling for seasonal and spatial variation, there was evidence of a 39 % longitudinal reduction in HBR (supplementary Table 2) from the beginning to end of the study period. Similarly, after controlling for variation due to season and village there was also evidence of 23% decline in the proportion of *An. coluzzii* relative to *An. gambiae* over the study period (Supplementary Table 2, Figure 2).

**Figure 2:**
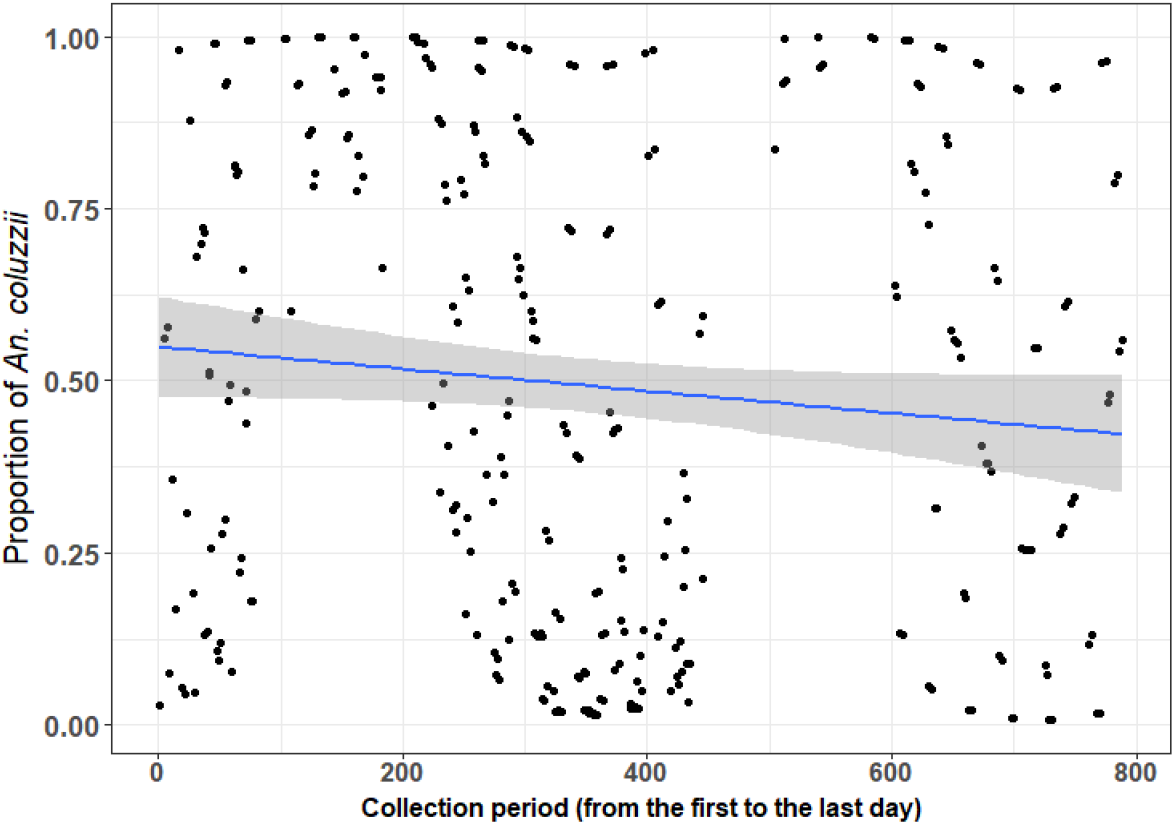
Mean predicted long-term trends in *Anopheles coluzzii* proportion within the *An. gambiae* s.l. complex from 12 villages in southwestern Burkina Faso. The blue curve and line represent the predicted regressions from models accounting for additional variation due to village and trapping location (inside and outdoors houses), with the grey-shaded area around them indicating the 95% confidence intervals.

### *Anopheles gambiae* s.l. behaviours (biting and resting) and host choice

Overall, *Anopheles gambiae* s.l. biting activity started very low earlier in the evening (7 – 8 pm) and steadily reached to a peak between 00 h and 04 h with the median biting time between 01 h and 02 h. Overall, about 54% [95% CI: ∼51 – 57%] of *An. gambiae* complex members were collected host-seeking outdoors. However, there was no long-term change in the biting location across the study (Supplementary Table 2). Further analysis of the subset of *An. gambiae* s.l. that were individually identified to species level indicated that the proportion of outdoor biting varied between *An. gambiae* and *An. coluzzii* (df = 1, χ2 = 6.82, p = 0.009). In addition, results here indicate that these vectors were less abundant in the indoor compare to the outdoor environment. Though, *An. gambiae* was somehow host-seeking more outdoor (54.73%, [95% CI: 52.35 - 57.12] compare to *An. coluzzii* [51.4%, 95% CI: 48.9 – 53.9%]).

The host-seeking activity showed there was similar patterns between the indoor and outdoor environment (Figure 3). Although, the peak of biting occurred around mid-night, some *An. gambiae* s.l. were caught still biting around 6 am (Figure 3). No longitudinal change in median biting time was observed (Supplementary Table 2). Further analysis of the subsample of *An. gambiae* s.l. identified to species level indicated there was difference in the median biting time between *An. coluzzii* and *An. gambiae (*Supplementary Table2). *An. coluzzii* was found to bite earlier (11 pm – 01 am) compared to *An. gambiae* (00 pm – 02 am). Additionally, the overall proportion of *An. gambiae* s.l. females resting outdoors was ∼46.97%. After controlling for the effect of season there was no evidence of a longer-term change in the proportion of female *An. gambiae* s.l. resting outdoors over the study period (df = 1, χ2 = 2.67, p = 0.32). Of the 94 *An. gambiae s*.*l*. females from which blood-meals could be identified, 52% were from human only, 35.11% were cattle only, and 12.77% contained a mixture of cattle and human blood (Table 3) resulting in an HBI of 64.9%.

**Table 3:**
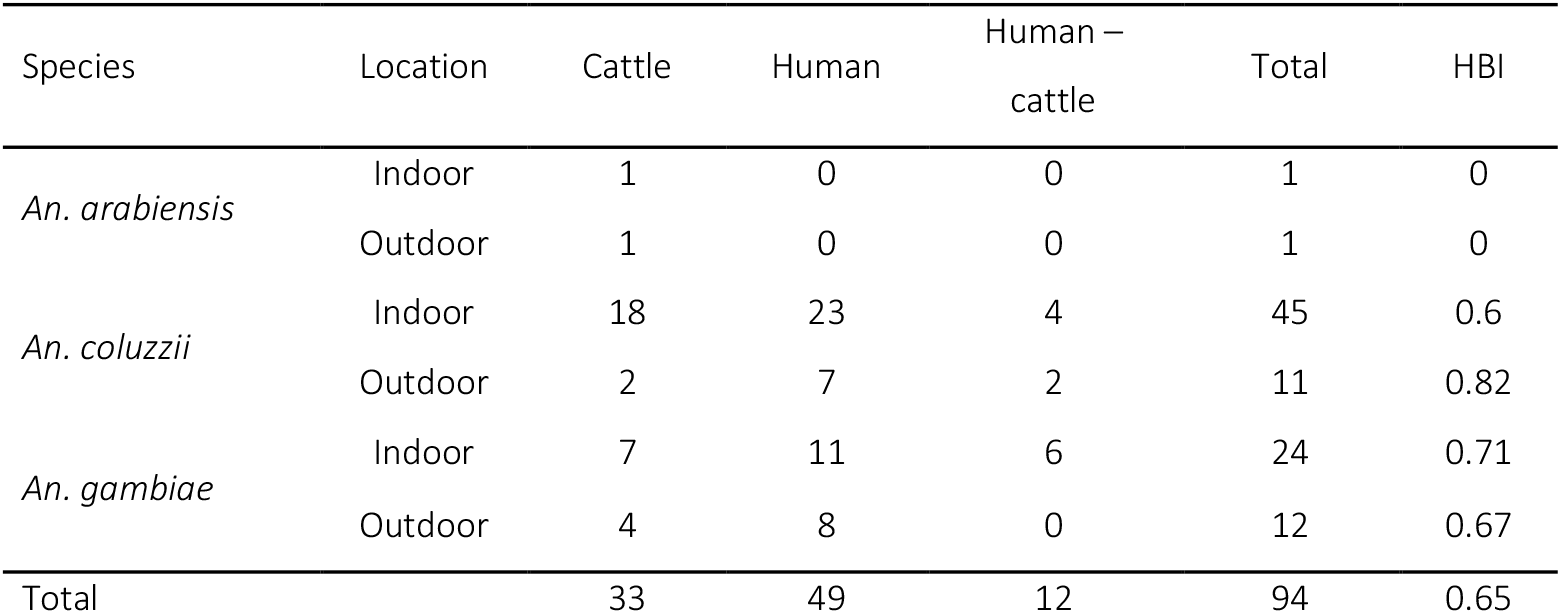
Total numbers of blood-fed female *An. gambiae* s.l. caught using Resting Bucket Traps in the 12 villages, from October 2016 to December 2018 (RBT) and display by species and trapping location pool over villages, according to the blood source. HBI indicate the proportion of mosquito that blood-fed on human.

| Species | Location | Cattle | Human | Human – cattle | Total | HBI |
| --- | --- | --- | --- | --- | --- | --- |
| *An. arabiensis* | Indoor | 1 | 0 | 0 | 1 | 0 |
|  | Outdoor | 1 | 0 | 0 | 1 | 0 |
| *An. coluzzii* | Indoor | 18 | 23 | 4 | 45 | 0.6 |
|  | Outdoor | 2 | 7 | 2 | 11 | 0.82 |
| *An. gambiae* | Indoor | 7 | 11 | 6 | 24 | 0.71 |
|  | Outdoor | 4 | 8 | 0 | 12 | 0.67 |
| Total |  | 33 | 49 | 12 | 94 | 0.65 |

**Figure 3:**
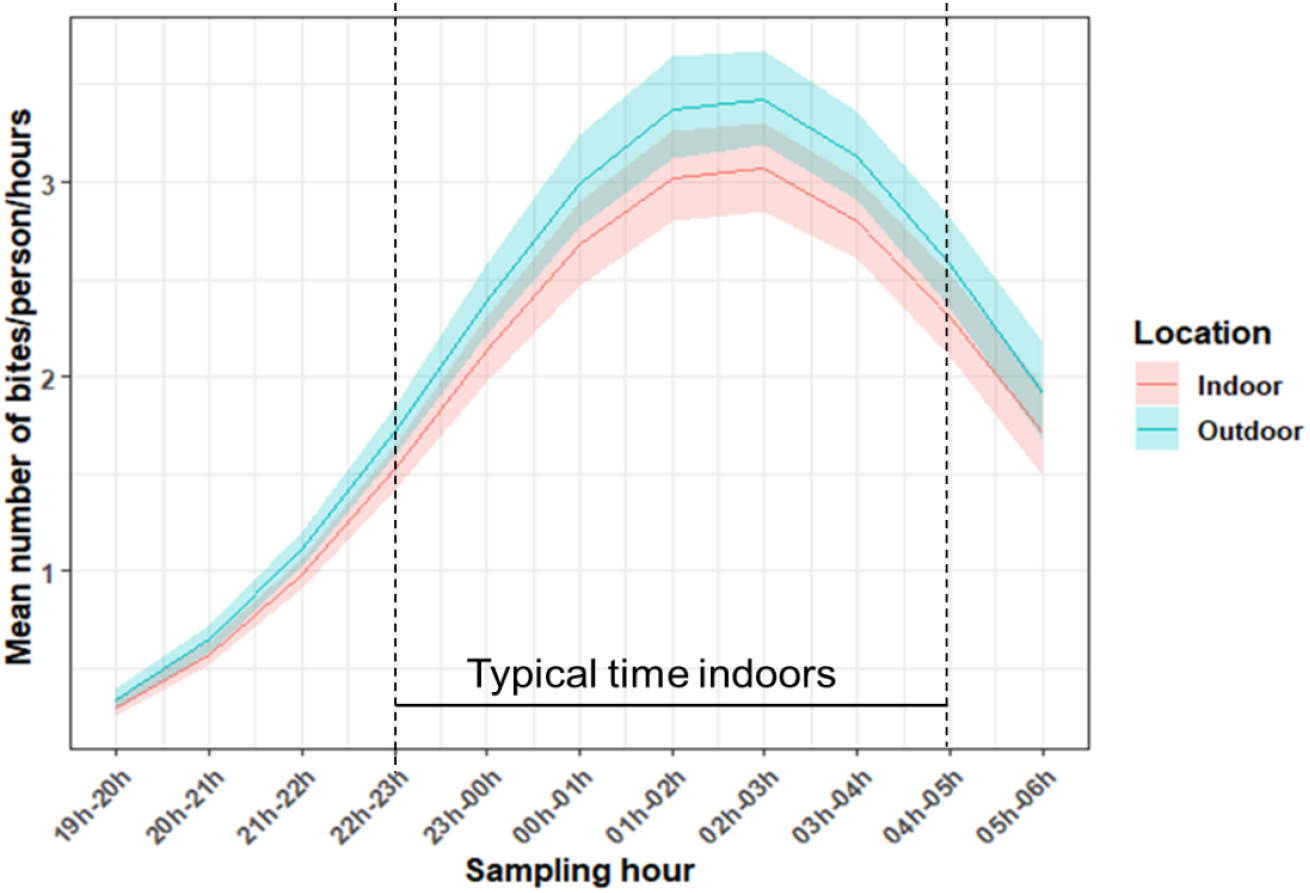
Mean number of *An. gambiae* s.l. biting per hour (as assessed from Human Landing Catches) in 12 villages in southwestern Burkina Faso, from October 2016 to December 2018. Data are pooled over village and collection period. The period between the red vertical dashed lines indicate period-time coinciding when most people are inside their dwellings. The blue and red full lines indicate the predicted number biting in outdoor and indoor settings, respectively; with the shaded areas around them indicating the 95% confidence intervals.

### Malaria transmission and LLIN effectiveness

Overall, 86.81% [95% CI: 83.6 – 90.02%] of *An. gambiae s*.*l*. was caught biting between 10 pm and 5 am (corresponding to time when most people are indoors, P_fl_). Furthermore, results showed that 85.45% [95% CI: 80.64 – 90.26%] of human exposure occurred when people are assumed to be indoors (πi). Thus, under the simplifying assumption that people remain under an LLIN between 10 pm -5 am [48], ∼85% of exposure to *An. gambiae* s.l. could be preventable by LLINs. Taking into account the variations due to village and season, results suggested that there was a modest but consistent decline (for ∼7%) in the proportion of biting taking place when people are supposed to be indoors (Supplementary Table 3, P_fl_: z = -3.14, p = 0.002, Figure 4a) over the study period. Similarly, there was also ∼10% decrease in the proportion of human exposure (Supplementary Table 3, πi: z = -3.72, p = 0.0002, Figure 4b).

**Figure 4:**
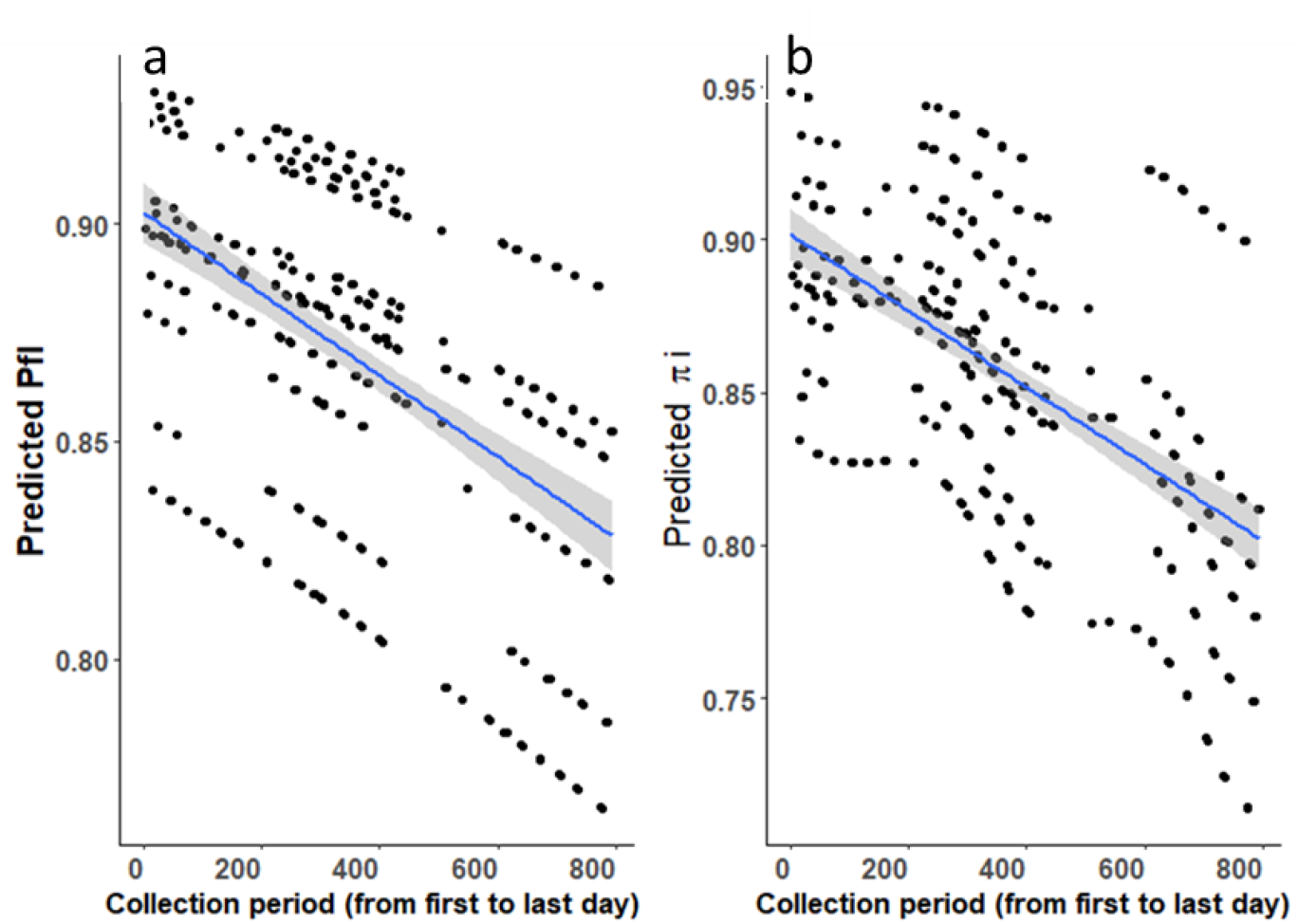
Predicted mean of a) Pfl = the proportion of *An. gambiae* s.l. bites occurring when most people are inside their dwellings and likely asleep (e.g. between 10 pm -5 am; Pfl), b) πi = total human exposure to *An. gambiae* s.l. bites occurring indoors (πi) based on Human Landing Catch from 6 villages over 2 years (Oct 1^st^, 2016 to Dec 4^th^, 2018) in southwestern Burkina Faso. Dots represents the predicted values (Pfl and πi) at each sampling night. The blue line represents the regression line from the model and the grey-shaded area the 95% confidence intervals.

Overall, in the study area, the mean sporozoite rate (SR) was 3.48% [95%CI: 1.51 – 5.26%] in *An. gambiae* s.l. Though, after taking into account, the seasonal and spatial variation, results suggested that the mean SR declined from ∼5% to ∼2% over the study period (z = -2.5, p = 0.01, Supplementary Table 4, Figure 5). However, no significant difference in this SR was found between species (*An. coluzzii* and *An. gambiae)* or host-seeking location (indoor versus outdoor, Supplementary Table 4). Further, the Entomological Inoculation Rate (EIR) was calculated to estimate the potential epidemiological impact of temporal changes in vector abundance and sporozoite rate. This was done based only on data from the subset villages (06) monitored in both year 1 (Oct 2016 -Sept 2017) and year 2 (Oct 2017-Sept 2018). Here, the annual EIR was considerably higher in year 1 (289.25 infective bites per person per year) compare to that from year 2 (94.81 infective bites per person per year). This decline was in accordance with that of the human biting rate and SR. Despite this decline, residents were still predicted to be exposed to 29 infective bites per person per year after adjustment for the proportion of bites (∼85%) that can be prevented through effective use of LLINs.

**Figure 5:**
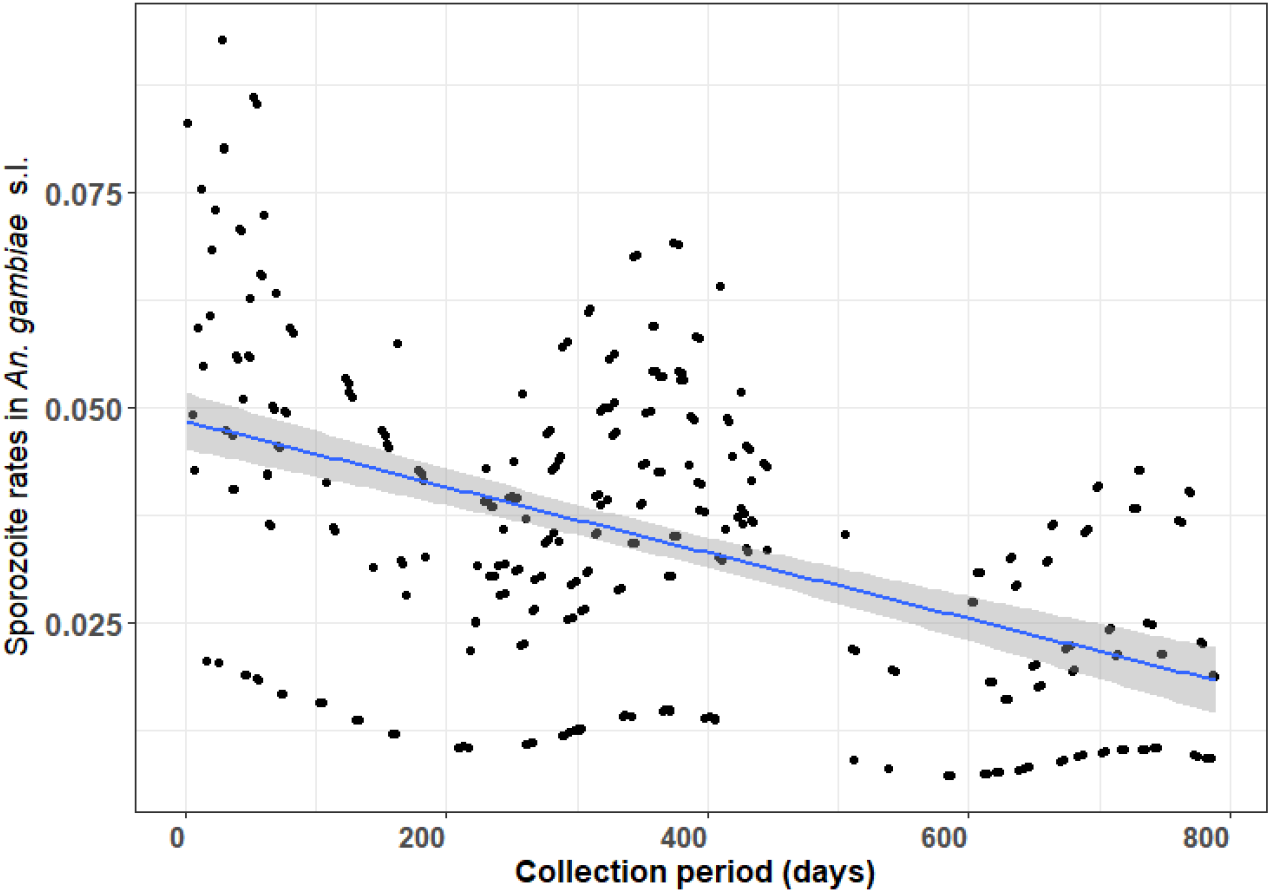
Mean predicted longer-term trend in sporozoite infection rates in *An. gambiae* s.l. (dots). Data are from 12 villages in southwestern Burkina Faso using Human Landing Catches from October 2016 to December 2018. The curve blue and line indicate the regression curve and line respectively and the grey-shaded areas around them indicate 95% confidence intervals.

## Discussion

Here we tested for evidence of systematic changes in insecticide and behavioural resistance traits in malaria vectors over 2.5 years following a mass LLIN distribution in Burkina Faso. Consistent with expectation, there was a high prevalence of insecticide resistance (IR) within these malaria vector populations, which further intensified over the period of the study, with post-exposure mortality falling by ∼ 50% (from 38% at the start to 17% at the end). In contrast, although baseline levels of “behavioural avoidance” traits were higher than expected (e.g. > 50% outdoor biting and resting), behavioural phenotypes were relatively stable across the study period. There was however a reduction in *An. gambiae* s.l. abundance (Human biting rate) and a shift in vector species composition across the study period; with a moderate reduction in the proportion of *An. coluzzii* relative to *An. gambiae*. The potential epidemiological impacts of these ecological changes are diverse. On one hand, there was a significant drop in the Entomological Inoculation Rate between years, indicating a fall in human exposure and malaria transmission. On the other, the proportion of exposure to *An. gambiae* s.l. expected to be preventable by LLIN use also fell over the study period (πi, ∼7% fall); indicating a moderate erosion of protection from residual transmission through time. We hypothesize that the fall in πi was due to a shift in vector species composition, with the slightly more exophilic *An. gambiae* increasing relative to *An. coluzzii* through time. Thus, we hypothesize that while both IR and avoidance behaviours are contributing to the limited impact of LLINs in this setting, behavioural traits appearing to be responding more slowly to selection from LLIN.

The rise in IR across this study is in line with similar increases observed over short time periods in other African settings [49, 50]. This acceleration in IR appears to be a relatively recent phenomenon; with the post-exposure mortality of *An. gambiae s*.l. to the DD in this setting being relatively similar in the 1970s when synthetic pyrethroid were first introduced for pest control (∼95%, [51]) to that reported in the 2010s [52]. The intensification of IR over the last decade is likely due to the initiation of mass LLIN distribution programmes in 2010. For example, post-exposure mortality of *An. gambiae s*.*l*. to the DD in one of our study sites (Tiefora) was ∼39% [53] in 2014 compared to only ∼15% reported here (2016-18). Additionally, in Tengrela (another study village), post exposure mortality to DD declined from ∼92% to ∼19% between 2011 and 2013 [37]. Considering the rapid rise in IR, the government of Burkina Faso has recently switched 2019 to new net products that combine pyrethroids with other insecticide classes (e.g. Interceptor G2: alpha-cypermethrin and chlorfenapyr; PBO-LLINs: deltamethrin and the synergist piperonyl butoxide).

While insecticide resistance increased significantly over the study, mosquito vector behaviours were generally static. However, some mosquito behavioural traits observed here differ substantially from historical reports in the study area; raising the possibility of more gradual long-term changes that may not be detectable over a few years. For example, previous studies from the Central, Plateau Central and West regions (2001-2015) of Burkina Faso reported *An. gambiae* s.l. was more likely to bite indoors than outdoors [43, 54], or had an even split between indoor and outdoor biting [46]. In contrast, here *An. gambiae* s.l. was more likely to host seek outdoors [∼54%; 95% CI: ∼51 – 57%]. Additionally, a recent review of *An. gambiae* s.l. biting behaviour from a range of African countries that concluded in general > 80% of vector bites occur indoors [28]; substantially higher than observed here. *Anopheles gambiae* s.l. were also found in indoor and outdoor resting traps with relatively similar frequency (53% versus 47%); in contrast to earlier studies in south western Burkina Faso where indoor resting was more common (∼67%; [55]). The Human Blood Index (∼65%) of these *An. gambiae* s.l. populations was also lower than previously reported in Burkina Faso (> 77%; [56]) and Benin (> 90%, [57]). Thus, although there was no clear evidence of mosquito behavioural change across the two years of this study, comparison with historical and wider regional data suggest that lower acting adaptations may be ongoing in tandem with more rapid insecticide resistance.

LLINs have also been hypothesized to generate selection on mosquito biting times; with vectors shifting to bite earlier in the evening before people go to bed [58] to avoid this intervention. Here, *An. gambiae* s.l. still exhibited the characteristic “late night” pattern of biting, with no evidence of a shift over the study. The biting pattern observed here is consistent with early work on *An. gambiae* s.l. before mass LLIN use which also found biting rates peaked around 00 h [12, 59]. Similarly, a study in western Kenya found *An. gambiae* s.l. and *An. funestus* continued biting late in the night even in presence of LLINs [60]. However, other studies have detected substantial shifts in malaria vector biting times in association with interventions, including a recent report from Senegal of *An. funestus*, biting earlier morning and into daylight hours [61]. Adaptations in biting time may arise more rapidly in other settings due to variation in local ecology and background levels of IR.

Some previous studies that have reported shifts in malaria vector behaviour (e.g.[27, 62]), did not identify to species level, thus preventing interpretation of whether changes were due to within species adaptations or ecological shifts in species composition. While behavioural phenotypes remained relatively fixed within vector species in this study, there was an evidence of a longitudinal shift in malaria vector species composition characterized by a reduction in *An. coluzzii* relative to *An. gambiae*. The proportion of outdoor biting was slightly higher in *An. gambiae* (55%) than *An. coluzzii* (51%), thus the relative decline in *An. coluzzii* is consistent with the prediction that LLINs will have a greater impact on endophagic vector species. For example, highly anthropophagic and endophagic vector species like *An. gambiae* [63] and *An. quadriannulatus* [64] have decline more significantly than the more zoophagic and exophagic *An. arabiensis* following ITN introduction in East Africa. This adds to the growing body of evidence from across Africa showing that LLINs can provoke shifts in malaria vector species communities. Such ecological shifts may occur more rapidly than longer-term behavioural adaptations within species.

Taken in combination, these results suggest that IR may be the first line of defence to ITNs compare to behavioural change in mosquito vectors given most human hosts are indoors, and/or that behavioural phenotypes have less capacity for adaptation than those involved with IR. Differences in the rate of change in IR and mosquito behavioural traits also likely reflect their genetic basis. Mosquito behaviours are likely complex multigenic traits [65], governed by many genes (some possibly antagonistic) that may limit capacity to respond quickly to selection. There is some evidence that behaviours like human host choice [65] and outdoor/indoor biting or resting [66] have a genetic basis in African malaria vectors. The genetic basis of IR traits is well established [18, 67], with some IR mechanisms linked to a single point mutation (e.g. Knock-down resistance [16] ; indicating this type of resistance may respond rapidly and efficiently to selection. It is known that vector population in Burkina Faso evolved high frequency of target site mutation (e.g. frequency of the L1014F type > 0.8 [68]) that may explain why they may be able to adapt quicker.

The potential epidemiological impacts of the vector adaptations observed here are complex. On one hand, results indicate that LLINs can still effectively prevent the bulk of human exposure to malaria vectors (∼85%) in this setting. Additionally, vector abundance, malaria infection rates and the associated Entomological Inoculation Rate fell across the study; suggesting a transmission decline. However, this degree of protection expected from LLIN use (85%) is somewhat lower than estimated from other parts of Burkina Faso (90%, in 2002-2004), and Africa (95 – 99%; [69]). Furthermore, the longitudinal changes in vector populations described here can further diminish LLLIN impact. First, the intensification of IR is likely to erode the community protection, afforded to non-net users. Second the proportion of exposure preventable by using LLINs (πi) was predicted to decline over the study period. Even if the proportion of human exposure preventable by LLIN use remained at current levels, high levels of residual transmission will persist because of the relatively high abundance and malaria infection rates in these vector populations. People in this area are estimated to be exposed to between ∼215 infective bites per person per year even if they sleep under an LLIN between 10 pm – 5 am. The small shifts in P_fl_ and πi observed through time could further increase this exposure. Interventions that tackle both insecticide resistant and outdoor biting mosquitoes will thus be needed to tackle residual transmission.

## Conclusions

Both insecticide resistance and behavioural avoidance strategies in *An. gambiae* s.l. are contributing to the erosion of malaria vector control impact across Africa. In a longitudinal study in Burkina Faso, we show that IR increases more rapidly than behavioural adaptations following the mass distribution of insecticidal nets. Although mosquito behaviours stayed relatively stable over the study, baseline traits indicated a higher capacity for ‘behavioural avoidance’ of nets (through outdoor biting and resting) than historical data. This highlights the possibility that behavioural traits are adapting but at a more gradual rate than physiological resistance. As most human exposure to infected mosquitoes still occurs indoors in this and other African settings, effective indoor interventions must still be prioritized including the use of novel insecticide classes on nets and sprays and non-insecticidal approaches. However, the proportion of exposure occurring outdoors is notable and increasing. Supplementary measures tackling human exposure in and outdoors will thus be required to reclaim progress and tackle residual transmission in this and other high burden African settings.

## Methods

### Study site

This study was conducted in 12 villages within the Banfora District, in the Cascades Region, south-western Burkina Faso (Supplementary Table 5, Figure 6). In 2018, the population size of the region was 822,445 [70]. The region has a humid savannah climate characterized by a rainy (May to October) and dry season (November to April). Annual rainfall ranges from 600-900 mm; with a mean temperature of ∼26 °C [15.7 °C -38.84 °C] and humidity of ∼62% [15.11 -99.95%] during the study. Malaria is endemic in the region, with an incidence of Plasmodium falciparum being ∼3 episodes per child of 6 -14 years old over the six month annual transmission season [32].

**Figure 6:**
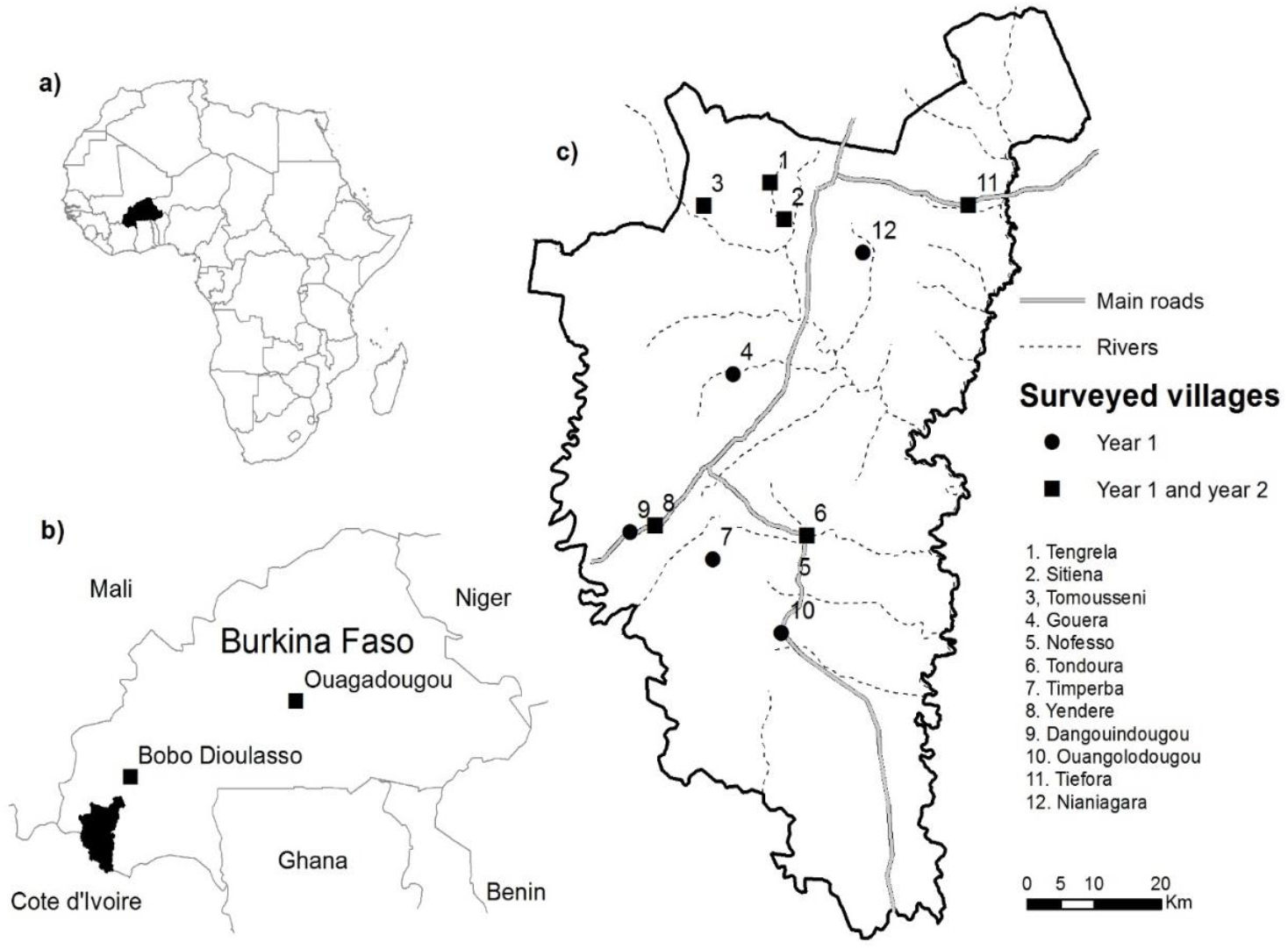
Map of the 12 study villages. a) location of Burkina Faso within Africa, b) study area in the Cascades Region in Burkina Faso, and c) villages where mosquito collection took place. Circles represent the villages sampled for 18 months and squares represent the villages that were part of the longer-term study site and where sampling was extended to 26 months.

### Insecticide resistance monitoring

We aimed to measure IR in all 12 original study sites at least once during the wet and dry season of each year. Due to the limited availability of larval habitats in some villages and months (especially during the dry season), IR monitoring was only possible in 9 villages (Supplementary Table 6 & 7). Larvae were collected from aquatic habitats and reared to adulthood under standard conditions [71]. Insecticide resistance assays were performed by exposing groups of 22 -27 adult female *An. gambiae s*.*l*. to the World Health Organization discriminating dose for deltamethrin (DD = 0.05%, a concentration that can kill 99.9% of a susceptible population [72]), 5 times (0.25%), 10 times (0.5%) and 15 times (0.75%) the DD respectively (Supplementary Table 6). Mosquitoes were exposed to insecticide-treated papers obtained from WHO / Vector Control Research Unit, University Sains Malaysia [73], following WHO guidelines for Tube bioassays [72].

### Assessment of human biting rate and behaviours

Vector surveillance was conducted to measure human biting rates (HBR) and mosquito resting behaviours bi-monthly at all 12 sites for the 16 months (Oct 2016 -Feb 2018, Figure 6). Additionally, surveillance continued for a further 10 months (Feb-Dec 2018) in a subset of 6 villages to generate longer-term data. The longer-term study villages were selected to achieve a relatively broad spatial distribution.

Host-seeking mosquitoes were sampled twice a month in each village from 2 different households per day using Human Landing Catches (HLC). Here, men collected mosquitoes landing on their exposed leg using a mouth aspirator and flash torches [74]. Collection took place both inside houses and outdoors (the peridomestic area) from 7 pm to 6am each night. During each hour of collection, participants caught mosquitoes for 45 minutes, followed by a 15-minute rest break. Participants involved in mosquito collections rotated between indoor and outdoor trapping stations each hour to avoid confounding location with individual differences in attractiveness to mosquitoes.

In addition, Resting Box Traps (RBTs) were placed in and outside houses [75] to sample resting mosquitoes. These RBTs were made locally using 20 L plastic buckets, with their inner surface covered with moistened black cotton cloth to create a high contrast and humid environment. On each night of collections, two RBTs were placed in the same households where HLCs took place (set within different houses in the compound). Inside houses, RBTs were placed on the floor in a relatively shaded corner of the sitting room. Two RBTs were also set outdoors at ∼8 metres from houses to capture outdoor resting mosquitoes. RBTs were set up at approximately 7 pm and emptied the following morning (∼5 am) using electrical aspirators.

### Mosquito processing

Mosquitoes collected in HLCs and RBTs were transferred to an insectary in Banfora town and sorted to species complex level using morphological keys [76]. Of those identified as belonging to the malaria vector group *An. gambiae* s.l., a subset of 7852 females (∼20 % of total) were selected to provide a representative sample from each month, village, trapping location (indoor *vs* outdoor) for identification to species level by PCR [77]. The subsampling strategy is described elsewhere [78]. Furthermore, malaria infection in mosquitoes was assessed by testing for the presence of *Plasmodium falciparum* circumsporozoite protein (CSP) in their head and thoraxes using a monoclonal sandwich Enzyme Linkage Immuno-Sorbent Assay (ELISA) developed by [79]. Additionally, female *An. gambiae* s.l. from the RBT collections were visually graded according to their repletion status (abdominal condition) into categories of blood-fed, unfed, gravid, and half gravid [80]. Blood-meal identification was carried out on blood-fed *An. gambiae* s.l. to determine the origin of their blood-meal (human, cattle or both [81]) using a direct Enzyme Linkage Immuno-Sorbent Assay (ELISA; [82]). The degree of “anthrophagy” in vectors was estimated in terms of the “human blood index” (HBI) [83], defined as the proportion of identified blood meals taken from humans. In ELISA tests (for sporozoite and blood meal source identification), two technical replicates of each sample were run in two different microplates at the same time and retested in cases where the first result was ambiguous. The absorbance of the solutions/reactions at the end of each ELISA was measured using microplate reader (Elx808; Bio-Tek) at 450 nm. To avoid any false positives (due to background noise), a sample was considered positive for an assay when its optical density (OD) was 2-fold higher than the average of the OD of both negative controls. Positive controls were also used in all ELISAs to ensure the procedure was working.

### Data analysis

Statistical analysis was conducted with the aim of testing for “longitudinal shifts” (over 26 months) in key indicators of vector demography, species composition, IR and behaviour using Generalized Additive Mixed-Models (GAMMs) in the R statistical software (version 3.6.1 of 2019-07-05) [84]. Insecticide resistance was measured in terms of mosquito mortality 24 hours after exposure to deltamethrin (different doses). Long-term trends in all ecological, behavioural and insecticide resistance variables were assessed after controlling for spatial (between-village) and environmental (season, temperature and humidity) variation. The key response variables were: (i) the proportion of mosquitoes dying within 24 hours of DD exposure, (ii) Human biting rate and species composition (proportion of *An. coluzzii* in *An. gambiae s*.*l*.), (iii) proportion of outdoor biting and resting, (iv) median biting time, (v) proportion of human exposure to bites, vi) the sporozoite rate and the vii) the entomological inoculation rates. To assess longer-term trends occurring over the 26 months of the study, a temporal variable was created by assigning a continuous value to each day of collections from the first (1 October 2016) until the last (4 December 2018), hereafter called “Longer_term” (Supplementary Table 8). This was incorporated into models as a continuous covariate to test for evidence of a consistent temporal rise or decline after accounting for seasonal and spatial variation. Seasonality was incorporated by fitting a smoothing spline where each day of the year was classified on a scale running from 1 (set as January 1^st^) to 365 (December 31^st^) hereafter called “Season” (Supplementary Table 8), and spatial variation by fitting village as a fixed effect. The mean nightly temperature and humidity at collection households were derived from hourly values (7 pm -6 am) obtained from data loggers and included as additional explanatory variables (Supplementary Table 8).

Data on the time and location of biting were combined for estimating two metrics of human exposure to bites from *An. gambiae* s.l.: the proportion of *An. gambiae* s.l. caught during hours when typically, ≥ 50% of people were indoors and likely to be in bed (P_fl_). Based on visual surveys described in [78] over 672 households, we estimated that > 50% of residents were indoors between 10 pm and 5 am consistently across the study period; although more recent, detailed anthropological investigation indicates time indoors may vary substantially between individuals and seasons and be lower assumed here [48]. The P_fl_ was estimated following [42] by dividing the total number of *An. gambiae s*.*l*. collected indoors and outdoors when ≥ 50% of people were indoors (10 pm to 5 am) by the total number collected that night. The proportion of human exposure to bites that occur indoors (π_i_) was then calculated by dividing the number of *An. gambiae* s.l. collected indoors between 10 pm – 5 am by itself plus those collected outdoors when >50% of residents were outdoors and thus unprotected by LLINs (between 7 – 10 pm and 5 – 6 am (Supplementary Table 8). Within the R statistical software [84] GAMMs within the ‘mgcv package’ [85] augmented with the lme4 package [86] known as GAMM4 were used to test for associations between IR, all vector ecological and behavioural metrics and explanatory variables of village, season, “longer-term” trend and environmental factors. Details of fixed and random effects and distribution are given in Supplementary Table 8. Due to the low number of mosquitoes caught in RBTs, it was not possible to test for spatial and long-term variation in the abundance of resting mosquitoes. Instead, the proportion of *An. gambiae* s.l. resting outdoors across the whole study period was assessed. Mosquito host choice was assessed in terms of the HBI, the proportion of blood-fed *An. gambiae* s.l. tested positive for human blood out of the total from which blood-meals were identified (N = 94). Furthermore, the EIR defined as the average number of infective bites a person (ibppn) would expect to receive from *An. gambiae* s.l. in a given location per year. This EIR was calculated as the product of the mean nightly human biting rate (*ma*) and vector sporozoite infection rates (SR) multiplied by 365 days [41] for the six villages that were monitored over two years.

### Ethical approval

Ethical clearance was obtained from the Ethical Committee for research in Health of the Ministry of Health of Burkina Faso (EC V3.0_CERS N°2016-09-097) and the Institutional Bioethical Committee of the local research institution (National Malaria Research and Training Centre, CNRFP) under EC V3.0_ N°2016-026/MS/SG/CNRFP/CIB) and the Liverpool School of Tropical Medicine (Certificate 16-038). Prior to starting the research, the project aims, and objectives were explained to community leaders in each village. Signed informed consent was also obtained from all household owners where mosquitoes were collected, and volunteers who took part in mosquito collections by HLC. We confirm that all methods were performed in accordance with the relevant guidelines and regulations.

## Data Availability

All data are provided as Figures and Supplementary materials

## Data accessibility

All data are provided as Figures and Supplementary materials

## Author’s contributions

AS designed the study, conducted the field data collection, did the molecular analysis, performed the data analysis, interpreted the results, and drafted the manuscript. WMG, NFS, HR and HMF contributed to the design of the study and the sampling protocol and provided comments upon the manuscript. HMF was a major contributor in writing this manuscript. MMT, FC and PO contributed to the molecular analysis in the labs. LN, JM, contributed to the data analysis. All authors read and approved the final version of the manuscript.

## Acknowledgements

We thank all the head of the villages, all the communities, household owners and the volunteers for their collaboration and help in data collection. Thank also to all the field and lab teams for the great job in the data collection and processing.

