## Supplementary material for "Insecticide resistance outpaces behavioural adaptation, as a response to Long-Lasting Insecticidal Net distribution, in malaria vectors in Burkina Faso"

**Supplementary Table 1:** Significance of explanatory variables included in the model for assessing variation in *An. gambiae* s.l. mortality rates. Here, df is the degree of freedom, Chi-sq ($\chi^2$) represents the values of Likelihood Ratio Test. and p indicates the p-values associated with each term. "Season" is a smoothing function on days from 1 - 365 describing a year of collection for assessing the seasonality in the proportion. "Longer-term" a discrete variable from first to the last day of collection (798) describing the long-term trend in the proportion.

| | 0.05% deltamethrin (Model 1) | | | 0.25% deltamethrin (Model 2) | | | 0.5% deltamethrin (Model 3) | | | 0.75% deltamethrin (Model 4) | | |
|---|---|---|---|---|---|---|---|---|---|---|---|---|
| Explanatory variable | Chi-sq | df | p | Chi-sq | df | p | Chi-sq | df | p | Chi-sq | df | p |
| Season | 15.1 | 3.95[a] | 0.003[b] | 1.54 | 0.7[a] | 0.12 | 0.0 | 0.0 | 0.31 | 0.0 | 0.0 | 0.13 |
| Longer-term | 20.91 | 1 | 0.00[bn] | 3.726 | 1 | 0.05[n] | 0.11 | 1 | 0.74 | 11.25 | 1 | 0.00[bn] |
| Village | 29.61 | 8 | 0.00[b] | 17.44 | 8 | 0.00[b] | 12.72 | 7 | 0.00[b] | 16.81 | 7 | 0.019[b] |

[a] indicates that the correspondent "df" represents the effective degree of freedom (edf); [b] indicates the significant terms in the model and [n] indicates negative therefore a long-term reduction in the mortality rate; P = 0.00 indicate p<0.001;

**Supplementary Table 2:** Output of the different models on proportion of *An. coluzzii*, the outdoor biting, the median biting time and the outdoor resting. Here, df is the degree of freedom, Chi-sq ($\chi^2$) represents the values of Likelihood Ratio Test. and p indicates the p-values associated with each term. “Season” is a smoothing function on days from 1 - 365 describing a year of collection, for assessing the seasonality in the proportion. “Longer-term” a discrete variable from first to the last day of collection (798) describing the long-term trend in the proportion.

| | Proportion of Anopheles coluzzii (Model 5) | | | Human biting rate (Model 6) | | | Proportion of outdoor biting (Model 7) | | | Median biting time (Model 10) | | |
|---|---|---|---|---|---|---|---|---|---|---|---|---|
| **Explanatory variable** | **Chi-sq** | **df** | **p** | **Chi-sq** | **df** | **p** | **Chi-sq** | **df** | **p** | **F** | **df** | **p** |
| Season | 68.35 | 5.22[a] | 0.04 | 6.63 | 1 | 0.01* | 3.06 | 1.55[a] | 0.19 | 89.66 | 1.92 [a] | 0.00 |
| Longer-term | 32.78 | 1 | 0.00[n] | 1165 | 6.84[a] | 0.0069* | 1.31 | 1 | 0.25 | 0.127 | 1 | 0.72 |
| Village | n/a | n/a | n/a | 230.54 | 11 | <0.0001* | 13.70 | 11 | 0.25 | 2.32 | 11 | 0.00 |
| Location | n/a | n/a | n/a | 21.28 | 1 | <0.0001* | n/a | n/a | n/a | 0.221 | 1 | 0.64 |
| Location: Village | 24.78 | 11 | 0.01 | 14.93 | 11 | 0.185 | n/a | n/a | n/a | 0.61 | 11 | 0.82 |
| Humidity | 0.04 | 1 | 0.84 | 0.004 | 1 | 0.95 | 2.76 | 1 | 0.09 | 0.21 | 1 | 0.65 |
| Temperature | 1.53 | 1 | 0.22 | 0.15 | 1 | 0.7 | 0.49 | 1 | 0.48 | 0.346 | 1 | 0.55 |
| Species | n/a | n/a | n/a | n/a | n/a | n/a | 6.82 | 1 | 0.009 | 23.47 | 1 | 0.00 |

[a] indicates that the correspondent “df” represents the effective degree of freedom (edf); [b] indicates the significant terms in the model and [n] indicates negative therefore a long-term reduction in the mortality rate; P = 0.00 indicate p<0.001;

**Supplementary Table 3** Output of the Model 14 and 15 on respectively the proportion of mosquito when people are indoor ($P_{fl}$) and human exposure to mosquito bite indoor ($\pi_i$). Here, df is the degree of freedom, Chi-sq ($\chi^2$) represents the values of Likelihood Ratio Test. and p indicates the p-values associated with each term. "Season" is a seasonal spline from 1 - 365 describing a year of collection, and "longer-term" a discrete variable from first to the last day of collection (798) describing a linear term representing longer-term linear change across the study period.

| | Proportion of mosquito when people are indoor ($P_{fl}$) (Model 14) | | | Human exposure to mosquito bite indoor ($\pi_i$) (Model 15) | | |
|---|---|---|---|---|---|---|
| **Explanatory variable** | **Chi-sq** | **df** | **p** | **F** | **df** | **p** |
| Season | 0.01 | 0.02 [a] | 0.39 | 1.42 | 0.86 [a] | 0.11 |
| Longer-term | 9.86 | 1 | 0.00[n] | 13.81 | 1 | 0.00[n] |
| Village | 50.16 | 11 | 0 | 41.51 | 11 | 0 |
| Humidity | 0.34 | 1 | 0.56 | 0.29 | 1 | 0.59 |
| Temperature | 1.28 | 1 | 0.126 | 3.58 | 1 | 0.06 |

[a] indicates that the correspondent "df" represents the effective degree of freedom (edf); [b] indicates the significant terms in the model; [n] and indicates negative therefore a long-term reduction in the mortality rate; P = 0.00 indicate p<0.001;

**Supplementary Table 4**: Significance of explanatory variables included in the Model 13 for *Plasmodium falciparum* sporozoite rates. Here, df is the degree of freedom and Chi.sq ($\chi^2$) represents the values of Likelihood Ratio Test; "Season" is a smoothing function on days from 1 - 365 describing a year of collection, for assessing the seasonality in the proportion. "Longer-term" a discrete variable from first to the last day of collection (798), describing the long-term trend in the proportion. The temperature and relative humidity were obtained by averaging the records over the course of the collection night.

| **Explanatory variable** | **Chi.sq** | **df** | **p-values** |
|---|---|---|---|
| Season | 3.175 | 1.3[a] | 0.036* |
| Humidity | 0.54 | 1 | 0.46 |
| Location | 0.02.49 | 1 | 0.12 |
| Location: Species | 1.02 | 1 | 0.31 |
| Longer-term | 6.26 | 1 | 0.01* |
| Species | 0.05 | 1 | 0.82 |
| Temperature | 2.41 | 1 | 0.12 |
| Village | 34.61 | 11 | 0.0002* |
| Village : Species | 3.35 | 11 | 0.95 |

* indicates the significant terms with p < 0.05 and [a] indicate the given df represent the estimate degree of freedom (edf = degree of wigginess) from smoother term.

**Supplementary Table 5:** Global Positioning System (GPS) coordinates and altitudes of the 12 study villages.

| Village | North | East | Altitude |
|---|---|---|---|
| Dangouindougou | 10° 11.970' | 005° 0.763' | 294 |
| Gouera | 10° 24.233' | 004° 52.418' | 306 |
| Nianiagara | 10° 33 42.4' | 004° 41.801' | 324 |
| Nofesso | 10° 08.226' | 004° 46.972' | 300 |
| Ouangolodougou | 10° 04.234' | 004° 48.329' | 307 |
| Sitiena | 10° 36.244' | 004° 40.244' | 286 |
| Tengrela | 10° 39.113' | 004° 49.465' | 285 |
| Tiefora | 10° 37.447' | 004° 33.201' | 302 |
| Timperba | 10° 09.897' | 004° 53.983' | 318 |
| Tondoura | 10° 11.792' | 004° 46.311' | 299 |
| Toumousseni | 10° 37.273' | 004° 54.893' | 274 |
| Yendere | 10° 12.473' | 004° 58.729' | 315 |

**Supplementary Table 6*:*** Number of females Anopheles gambiae s.l. exposed to different concentrations of deltamethrin and untreated paper (control) in bioassays between September 2016, December 2018 and summed across replicates. Numbers inside brackets are the total number of bioassay replicates conducted. "n/a" indicates no bioassays done at this level.

| | Concentrations of deltamethrin used | | | | | |
|---|---|---|---|---|---|---|
| Village | 0 | 0.05 | 0.25 | 0.5 | 0.75 | Total |
| Dangouindougou | 80 (3) | 200(8) | 50(2) | 51(2) | 49(2) | 430 |
| Gouera | 150 (6) | 54(2) | 148(6) | 125(5) | 72(3) | 549 |
| Nianiagara | n/a | 71(3) | n/a | n/a | n/a | 91 |
| Sitiena | 315(13) | 382(15) | 252(10) | 128(5) | 27(1) | 1,104 |
| Tengrela | 757(31) | 411(16) | 502(20) | 503(20) | 421(17) | 2,594 |
| Tiefora | 528(21) | 492(20) | 488(20) | 434(17) | 231 (9) | 2,173 |
| Tondoura | 282(11) | 300(12) | 240(10) | 147(6) | 123(5) | 1,092 |
| Toumousseni | 356(14) | 356(14) | 315(13) | 292(12) | 250(10) | 1,569 |
| Yendere | 205(8) | 253(10) | 167(7) | 186(7) | 71(3) | 857 |
| Total | 2,693 | 2,519 | 2,162 | 1,866 | 1,244 | 10,484 |

**Supplementary Table 7:** Number of females *Anopheles gambiae s.l.* exposed at different seasons to deltamethrin and untreated paper (control) in bioassays between September 2016 and December 2018, summed across replicates. Numbers inside brackets are the total number bioassay of replicates conducted. “n/a” indicates no bioassays done at this level.

| | Seasons | | | | | |
|---|---|---|---|---|---|---|
| Village | wet-16 | dry-17 | wet-17 | dry-18 | wet-18 | Total |
| Dangouindougou | n/a | n/a | 98(4) | n/a | 332(13) | 430 |
| Gouera | n/a | n/a | 549(22) | n/a | n/a | 549 |
| Nianiagara | n/a | n/a | 91(3) | n/a | n/a | 91 |
| Sitiena | 232(9) | n/a | 465(17) | 168(7) | 239(10) | 1,104 |
| Tengrela | 307(12) | 447(18) | 663(27) | 173(7) | 1,004(40) | 2,594 |
| Tiefora | 116(5) | 426(17) | 527(21) | 469(19) | 635(25) | 2,173 |
| Tondoura | n/a | n/a | 419(17) | n/a | 673(27) | 1,092 |
| Toumousseni | 464(17) | n/a | 371(15) | n/a | 734(29) | 1,569 |
| Yendere | 249(10) | n/a | 77(3) | 286(11) | 270(11) | 857 |
| Total | 1,368 | 873 | 3,260 | 1,096 | 3,887 | 10,484 |

**Supplementary Table 8:** Maximal models used for modelling seasonality including the primary response variable, explanatory variables and statistical distribution used. Hyphens (-) indicates no random effect used. Physiology means the abdominal status. "Longer_term" a discrete variable from first to the last day of collection (798) indicates the longer-term trend over the collection periods fitted as a linear term. The seasonality term was fitted using a non-linear smoothing function (spline $t_2$(Season, bs=cc)) on days as a period of 365-days. nHour and the I(nHour^2) respectively represent here hours as discrete variables from 1 being the first hour of collection (7pm-8pm) to the last hour of collection of the night being 11 (5am – 6am) and its quadratic term. Here, "subset of *An. gambiae s.l.* "refers to subset that were individually identified to species levels.

| Model | Trait | Response variables | Fixed Effect variables | Random effects | Type of data | Distribution |
|---|---|---|---|---|---|---|
| 1 | Mortality rate | (Dead/ (Dead +Alive)) | Village+ longer_term + $t_2$(Season, bs=cc), | Replicate | *Subset of An. gambiae* s.l. exposed to 0.05% of deltamethrin | binomial |
| 2 | Mortality rate | (Dead/ (Dead +Alive)) | Village+ longer_term + $t_2$(Season, bs=cc), | Replicate | *Subset of An. gambiae* s.l. exposed to 0.25% of deltamethrin | binomial |
| 3 | Mortality rate | (Dead/ (Dead +Alive)) | Village+ longer_term + $t_2$(Season, bs=cc), | Replicate | *Subset of An. gambiae* s.l. exposed to 0.5% of deltamethrin | binomial |
| 4 | Mortality rate | (Dead/ (Dead +Alive)) | Village+ longer_term + $t_2$(Season, bs=cc), | Replicate | *Subset of An. gambiae* s.l. exposed to 0.75% of deltamethrin | binomial |
| 5 | Proportion of *An. coluzzii* | (*An. coluzzii*/*An. coluzzii* + *An. gambiae*) | Village+ Location+ Temperature+ Humidity +Village : Location + Longer_term + $t_2$(Season) | Compound+ Household | Subset: *An. gambiae* s.l. lab-processed data | Binomial |

| Model | Trait | Response variables | Fixed Effect variables | Random effects | Type of data | Distribution |
|---|---|---|---|---|---|---|
| 6 | Human biting rate (HBR) | Number of *An. gambiae* s.l. | Village + Location + Temperature + Humidity + Year + Village: Year + $t_2$(Season, bs=cc) | Compound+ Household | Host-seeking nightly *An. gambiae* s.l. data from 6 villages | Negative binomial |
| 7 | Outdoor biting proportion | (Outdoor/ Outdoor +Indoor) | Village+ Temperature+ Humidity + Longer_term+ t2(Season, bs=cc), | Compound+ Household | Host-seeking nightly *An. gambiae* s.l. | Binomial |
| 8 | Outdoor biting proportion | (Outdoor/ Outdoor +Indoor) | Village+ Species+ Temperature+ Humidity + Longer_term+ $t_2$(Season, bs=cc), | Compound+ Household | Subset of *An. gambiae* s.l. identified to species level | Binomial |
| 9 | Mean number of bite/person/hours | Hourly count | nHour + I(nHour^2) | - | Host-seeking hourly *An. gambiae* s.l. | Negative binomial |
| 10 | Median biting time | Median of hours | Village+ Location+ Longer_term + Temperature + Humidity + Village : Location + $t_2$(Season, bs=cc) | Compound+ Household | Host-seeking hourly *An. gambiae* s.l. | Poisson |
| 11 | Median biting time by species | Median of hours | Species+ Village+ Location+ Longer_term + Temperature + Humidity + $t_2$(Season, bs=cc) | Compound+ Household | Subset of *An. gambiae* s.l. data | Poisson |
| 12 | Outdoor resting proportion (female) | (Outdoor/ Outdoor +Indoor) | Longer_term+ Species + Physiology + Species: Physiology +$t_2$(Season, bs=cc) | Village + Compound+ Household | Subset: *An. gambiae* s.l. resting lab-processed data | Binomial |
| 13 | Sporozoite rate | (Positive/Positive+ Negative)) | Village + Location + Species + Village: Species + Location: Species + Temperature + Humidity + Longer_term + $t_2$(cDate, bs = cc), | Compound+ Household | Subset of *An. gambiae* s.l. lab-processed from 6 villages | Binomial |
| 14 | Proportion of mosquito when people are indoor ($P_{fI}$) | $(I_{10pm\text{->}5am} + O_{10pm\text{->}5am})$ / $(I_{7pm\text{->}6am} + O_{7pm\text{->}6am})$ | Village+ Temperature + Humidity + Longer_term + $t_2$(Season, bs=cc), | Compound+ Household | Host-seeking nightly *An. gambiae* s.l. | Binomial |
| 15 | Human exposure to mosquito bite indoor ($\pi_i$) | $I_{10pm\text{ -> }5am}$ / $(I_{10pm\text{ -> }5am}$ + $O_{7pm\text{ -> }10pm,\ 5am\text{ -> }6am})$ | Village+ Temperature+ Humidity + Longer_term + $t_2$(Season, bs=cc), | Compound+ Household | Host-seeking nightly *An. gambiae* s.l. | Binomial |
